# Stroke burden in the Philippines (1990–2023): Temporal trends and risk factors from the GBD study

**DOI:** 10.64898/2026.03.04.26347658

**Authors:** Judah Israel Ong Lescano, Keith Pardillada Belangoy, Yoshito Nishimura, Ko Harada, Hideharu Hagiya, Quynh Thi Vu, Hanane Ouddoud, Gerard Lee See, Florencio V. Arce, Elizabeth Yu Tan, Naohiro Iwata, Tatsuaki Takeda, Yoshito Zamami, Toshihiro Koyama

## Abstract

**Background:** Stroke is a leading cause of mortality and disability globally. However, information about stroke burden in the Philippines is limited. We sought to analyze stroke burden in the Philippines from 1990 to 2023.

**Methods:** Incidence, prevalence, mortality, and disability-adjusted life-years (DALYs) estimates from the Global Burden of Disease Study 2023 data were used as indicators to analyze the burden of stroke by sex and age. Temporal trends in both crude and age-standardized rates were analyzed using joinpoint regression analysis.

**Results:** In 2023, stroke incidence was estimated at 156.2 (95% uncertainty interval [UI]: 140.8–175.4) thousand, prevalence at 1.2 (95% UI: 1.2–1.4) million, mortality at 72.2 (95% UI: 63.2–83.0) thousand, and DALYs at 2.1 (95% UI: 1.8–2.3) million. High systolic blood pressure was the leading contributor to risk-attributable stroke mortality and DALYs. Since 1990, age-standardized rates declined significantly, whereas crude rates increased markedly. Compared with women, men had a higher fatal burden and consistently exhibited a higher age-standardized burden. Although older adults (≥ 55 years) had the highest stroke burden and achieved reductions in stroke incidence and fatal outcomes, both fatal and non-fatal burdens consistently increased among young adults (35–54 years).

**Conclusion:** While age-standardized rates have improved, the rising crude burden and shift towards younger adults present significant public health challenges. These trends highlight the pressing need for aggressive and targeted risk factor control, sustained risk monitoring, and strengthened acute and post-stroke care to mitigate the growing health burden of stroke in the Philippines.

## Introduction

Stroke is a leading cause of mortality and long-term disability worldwide. In 2023, it was estimated to account for 11.31% of global death, ranking second among the leading cause, and 5.59% of global disabilities, assessed as disability-adjusted life years (DALYs), ranking third.^1^ Clinically, stroke is broadly classified into two types based on its etiology. Ischemic stroke occurs due to obstruction of cerebral blood flow, whereas hemorrhagic stroke, further classified into intracerebral or subarachnoid hemorrhagic stroke, is caused by rupture of cerebral blood vessels. Globally, although ischemic stroke accounts for most cases, hemorrhagic stroke is associated with a higher fatal burden.^2^

Reducing the burden of stroke requires a multi-pronged approach that focuses on preventing primary modifiable factors, surveillance and risk factor monitoring, acute and long-term care, financial protection, and health policy actions.^3^ However, the stroke system of care in the Philippines suffers systemic deficiencies characterized by insufficient number and disproportionate distribution of stroke specialists, acute stroke facilities, and post-stroke rehabilitation facilities. It is further compounded by uneven implementation of public health interventions toward stroke awareness and prevention, high out-of-pocket expenses, and poor control of modifiable risk factors such as hypertension, diabetes, and smoking.^3,4^

The recent data from the Global Burden of Disease (GBD) Study 2023 ranked stroke as the second leading cause of death and long-term disability in the Philippines^1^ While epidemiologic data on stroke in the Philippines have been previously reported^5,6^ most were published more than five years ago and variations in measuring methods and data sources hinder trend analysis and comparisons within the country and across global regions. Furthermore, no published studies have comprehensively analyzed the burden of stroke and risk-attributable stroke across four standard epidemiologic measures (incidence, prevalence, deaths, and DALYs), highlighting the need for a systematic assessment of the country’s stroke burden.

Therefore, this study aimed to analyze the burden of stroke in the Philippines at both the national and subnational level by age and sex from 1990 to 2023 using the GBD 2023 dataset. These findings will significantly improve our understanding of the burden of stroke in the country and provide valuable insights to policymakers and relevant stakeholders to help them develop and implement effective care services to affected and vulnerable populations.

## Methods

### Overview

The GBD 2023 study provides the most recent estimates across multiple epidemiologic measures and risk-attributable burden on 375 diseases across 204 countries and territories and 660 subnational locations from 1990 to 2023. These data (made available in October 2025) are publicly available at the Institute for Health Metrics and Evaluation website^1^ While the primary GBD study was conducted following the Guidelines on Accurate and Transparent Health Estimate Reporting, this secondary analysis was conducted in compliance with the Strengthening the Reporting of Observational Studies in Epidemiology guidelines. In this study, we used the data from the GBD 2023 study to estimate the trends in the burden of stroke and its pathological subtypes in the Philippines and its provinces in four standard epidemiological measures: incidence, prevalence, deaths, and DALYs. DALYs represent the combined measures of cause-specific mortality and non-fatal health loss, calculated by adding years of life lost (YLLs) and years lived with disability (YLDs) for each age-sex-location^7^

### Disease Definition

The 375 diseases and injuries included in the GBD 2023 study were organized into a four-level cause hierarchy, where stroke was classified as a third-level cause, while its subtypes were categorized as fourth-level cause. The reference case definition of stroke as well as its subtypes were according to the World Health Organization (WHO) and the International Statistical Classification of Diseases and Related Health Problems Tenth Revision (ICD-10) criteria^8^ The codes for ischemic stroke were ICD-10: I63–I63.9, I65–I66.9, I67.2–I67.3, I67.5–I67.6, and I69.3. The ICD-10 codes I61-I62, I62.1-I62.9, I68.1-I68.2, and I69.1-I69.2 were used to identify intracerebral hemorrhagic stroke while ICD-10 codes I60–I60.9, I62.0, I67.0–I67.1, and I69.0 were used to identify subarachnoid hemorrhagic stroke.

### Estimates of stroke

The GBD 2023 study added new studies that were not included in the previous systematic review for stroke models (GBD 2021). All estimates were based on vital registrations and both published and unpublished hospital records in the Philippines. However, data were excluded for locations where data points were implausibly low. A standard Cause of Death Ensemble model (CODEm) was used to model cause of death estimates for stroke. CODEm is a structured framework that runs multiple models on the same dataset and selects an ensemble that estimates death by location, age, sex, and year^2^ For non-fatal stroke modeling, DisMod-MR 2.1 (Seattle, WA, USA), a Bayesian modeling tool, was used to estimate stroke incidence, prevalence, YLLs, and YLDs stratified by location, age, and sex. These modeling frameworks account for missing data by utilizing covariates and spatiotemporal Gaussian process regression (ST-GPR). The detailed process has been described in previous studies^1,2^

### Risk factor estimation

The GBD 2023 study included 88 risks organized into four hierarchical levels at which risk combinations were evaluated at each level. Risk-attributable burden was estimated using a Comparative Risk Assessment framework that calculated the population attributable fraction relative to a theoretical minimum-risk exposure level (TMREL). TMREL represents the exposure level that results in the lowest possible health risk for a given population. The relationship (summary effect size) for each risk-outcome pair was estimated by either conducting a meta-analysis using the meta-regression–Bayesian, regularized, trimmed tool or by conducting a primary analysis of relationships with cause-specific mortality. Exposure was estimated using either a Bayesian meta-regression model (DisMod-MR 2.1) or a ST-GPR model. These approaches were used to pool data from various sources, control and adjust for bias in exposure data, and incorporate other information like country-level covariates. Furthermore, the model used a multiplicative function when aggregating the stroke burden across clusters and adjusted for mediation to prevent double counting of risk effects. Estimates are only given for deaths, YLLs, YLDs, and DALYs, and uncertainty intervals (UIs) were calculated using 250 draws. In this study, the risk factors analyzed were based on the ranked list of stroke mortality- and DALYs-related factors for 2023. The visualization is accessible through GBD Compare (http://vizhub.healthdata.org/gbd-compare).

### Statistical analysis

Temporal trends in both crude and age-standardized rates were analyzed through joinpoint regression analysis (Joinpoint Regression Program Software, Bethesda, MD, USA). Log transformation of rates was conducted to achieve a normal distribution. A permutation test of 4,499 permutations and an overall significance level of 0.05 were the parameters of the model selected to determine the optimal number of joinpoints^9^ The annual percent change (APC) for each segment and their weighted average (AAPC) was calculated, and their corresponding 95% confidence interval (CI) was estimated using the Emperical Quantile method with 5,001 resamples^10^ APCs and AAPCs were compared to zero and were considered significant if *p* was < 0.05 and remained stable. Figures in this study were generated using RStudio (Boston, MA, USA).

### Ethics consideration

Routinely collected, publicly available data was used in this study and therefore ethical approval, and informed consent was not required.

## Results

### Stroke Burden in 2023

In 2023, there were 156.2 (95% UI: 140.8–175.4) thousand new stroke cases, 1.2 (95% UI: 1.2–1.4) million prevalent cases, 72.2 (95% UI: 63.2–83.0) thousand stroke-related deaths, and 2.1 (95% UI: 1.8–2.4) thousand stroke-related DALYs in the Philippines. Although ischemic stroke was both the most incident (50.75%) and most prevalent (62.97%) subtype, intracerebral hemorrhage accounted for most stroke-related mortality (66.35%) and DALYs (70.89%). Analysis by sex revealed that age-standardized rates for ischemic stroke and intracerebral hemorrhage were consistently higher among men than among women. (Supplementary Tables S1–4). Stroke incidence, prevalence, mortality, and DALYs rates increased noticeably with age. Over half of new and existing stroke cases occurred among individuals aged 70 years and older, who also accounted for most stroke-related deaths and DALYs (Supplementary Tables S5–8). The proportion of stroke-related DALYs relative to total DALYs from all causes occurred in the 75–79-year age group (12.18%), whereas peak proportions for ischemic stroke and intracerebral hemorrhagic stroke were observed in the 85–89-year (7.57%) and 55–59-year (7.26%) age groups, respectively (Supplemental Table S9).

**Figure 1** shows a heterogenous distribution in age-standardized mortality and DALY rates at the provincial level in 2023. Davao Occidental had the highest estimated age-standardized DALYs (5,436.5 [4,529.7–6,392.0] per 100,000 population), followed by Sarangani (4,524.4 [3,394.1–5,339.3] per 100,000 population) and Davao Del Sur (3,695.9 [3,271.8–4,187.3] population). A similar pattern was observed for age-standardized stroke-related deaths, where Davao Occidental recorded 236.6 (95% UI: 194.4–281.7) per 100,000 deaths (Supplementary Tables S10–11).

**Figure 1.**
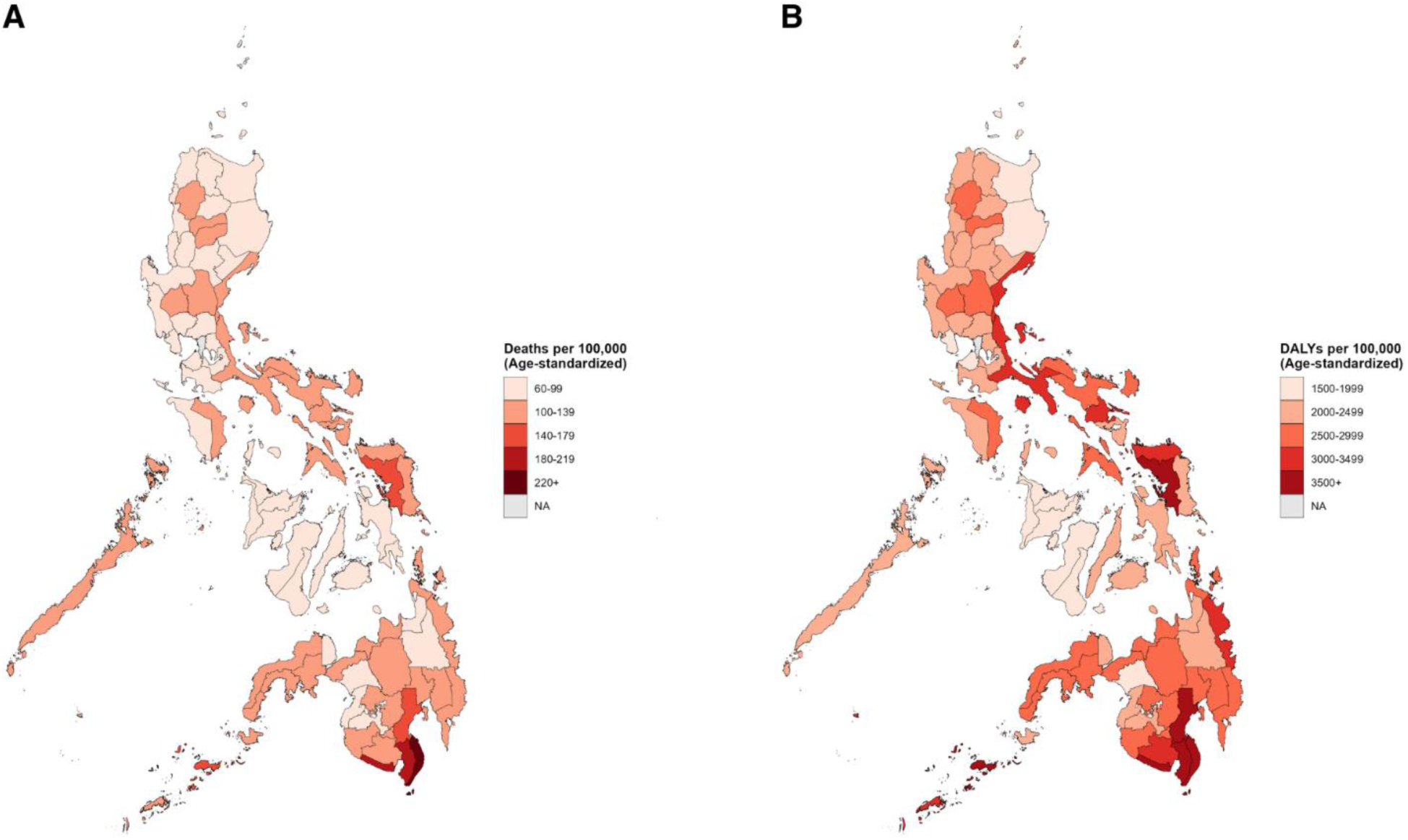
Crude and age-standardized mortality and disability-adjusted life years rates of stroke at provincial levels in the Philippines in 2023. (A) Age-standardized mortality (death) rate in 2023. (B) Age-standardized DALY rate in 2023.

### Risk-attributable stroke

**Figure 2** presents the ranking of risk-attributable age-standardized DALYs for stroke across the provinces with the highest DALYs in 2023, highlighting high systolic blood pressure as the leading contributor (1,285.1 [95% UI: 789.3–1,755.7] per 100,000 population), followed by household air pollution from solid fuels (386.9 [95% UI: 205.1.586.7] per 100,000 population) and smoking (356.6 [95% UI: 262.8–444.8] per 100,000 population), representing 66.43%, 19.91%, and 18.43%, respectively, of stroke DALYs attributed to all risk factors. For risk-attributable stroke-related deaths, kidney dysfunction ranked third ahead of smoking. Sex-specific analysis indicated that elevated low-density lipoprotein levels are a major contributor to stroke-related DALYs among women. All the risk factors for stroke contribute more significantly to both crude and age-standardized mortality and DALY rates of intracerebral hemorrhage stroke than of ischemic stroke (Supplemental Table S11–14).

**Figure 2.**
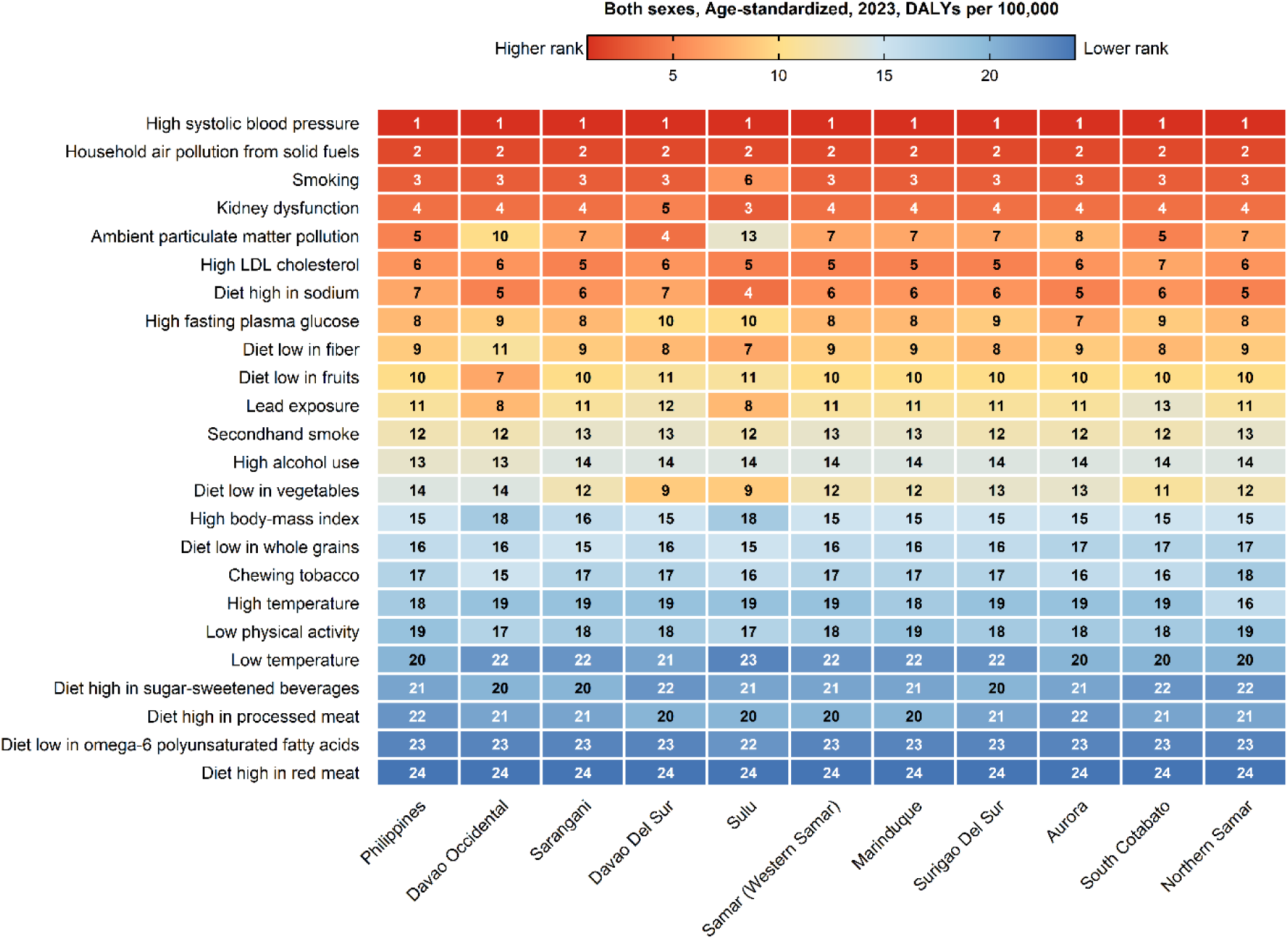
Risk factors of stroke in the Philippines

### Joinpoint analysis

Crude stroke mortality and DALY rates showed significant increases, as shown in **Table 1**, with an AAPC of 0.99% (95% CI: 0.76–1.16) and 1.05% (95% CI: 0.87–1.21), respectively. Among its subtypes, subarachnoid hemorrhage had the highest increase in AAPC across all measures (incidence: 1.18 [95% CI: 1.17–1.20]; prevalence: 1.42 [95% CI: 1.41–1.43]; mortality: 1.93% (1.72–2.12); DALYs: 1.79% [95% CI: 1.69–1.89]). However, stroke age-standardized mortality and DALY rates decreased significantly with an AAPC of −0.41% (95% CI: −0.55 to −0.28) and −0.36% (95% CI: −0.58 to −0.21), respectively. The AAPCs for ischemic and intracranial hemorrhagic strokes showed similar trends, whereas subarachnoid hemorrhagic stroke maintained a significantly increasing trend (Supplementary Figures S1–2).

**Table 1.** Average annual percentage change (AAPCs) of stroke from 1990 to 2023.

| Measure | Cause | AAPC | [95% Confidence Interval] |  | P-value |
| --- | --- | --- | --- | --- | --- |
| Crude |  |  |  |  |  |
| Incidence | Stroke | 0.93 | 0.91 | 0.95 | <0.001 |
|  | Ischemic Stroke | 1.02 | 1.01 | 1.04 | <0.001 |
|  | Intracerebral | 0.75 | 0.72 | 0.77 | <0.001 |
|  | Hemorrhage |  |  |  |  |
|  | Subarachnoid | 1.18 | 1.17 | 1.20 | <0.001 |
|  | Hemorrhage |  |  |  |  |
| Prevalence | Stroke | 0.96 | 0.95 | 0.97 | <0.001 |
|  | Ischemic Stroke | 1.12 | 1.10 | 1.14 | <0.001 |
|  | Intracerebral | 0.42 | 0.41 | 0.43 | <0.001 |
|  | Hemorrhage |  |  |  |  |
|  | Subarachnoid | 1.42 | 1.41 | 1.43 | <0.001 |
|  | Hemorrhage |  |  |  |  |
| Mortality | Stroke | 0.99 | 0.76 | 1.16 | <0.001 |
|  | Ischemic Stroke | 0.69 | 0.45 | 0.90 | <0.001 |
|  | Intracerebral | 1.06 | 0.86 | 1.23 | <0.001 |
|  | Hemorrhage |  |  |  |  |
|  | Subarachnoid | 1.93 | 1.72 | 2.12 | <0.001 |
|  | Hemorrhage |  |  |  |  |
| DALYs | Stroke | 1.05 | 0.87 | 1.21 | <0.001 |
|  | Ischemic Stroke | 0.94 | 0.76 | 1.08 | <0.001 |
|  | Intracerebral | 1.05 | 0.87 | 1.24 | <0.001 |
|  | Hemorrhage |  |  |  |  |
|  | Subarachnoid | 1.79 | 1.69 | 1.89 | <0.001 |
|  | Hemorrhage |  |  |  |  |
| Age-standardized |  |  |  |  |  |
| Incidence | Stroke | -0.37 | -0.39 | -0.35 | <0.001 |
|  | Ischemic Stroke | -0.34 | -0.36 | -0.33 | <0.001 |
|  | Intracerebral | -0.53 | -0.54 | -0.51 | <0.001 |
|  | Hemorrhage |  |  |  |  |
|  | Subarachnoid | 0.25 | 0.24 | 0.27 | <0.001 |
|  | Hemorrhage |  |  |  |  |
| Prevalence | Stroke | -0.07 | -0.08 | -0.07 | <0.001 |
|  | Ischemic Stroke | -0.10 | -0.11 | -0.09 | <0.001 |
|  | Intracerebral | -0.30 | -0.31 | -0.29 | <0.001 |
|  | Hemorrhage |  |  |  |  |
|  | Subarachnoid | 0.51 | 0.49 | 0.52 | <0.001 |
|  | Hemorrhage |  |  |  |  |
| Mortality | Stroke | -0.41 | -0.55 | -0.28 | <0.001 |
|  | Ischemic Stroke | -0.73 | -0.90 | -0.56 | <0.001 |
|  | Intracerebral | -0.52 | -0.75 | -0.33 | 0.006 |
|  | Hemorrhage |  |  |  |  |
|  | Subarachnoid | 0.63 | 0.40 | 0.86 | <0.00 |
|  | Hemorrhage |  |  |  |  |
| DALYs | Stroke | -0.36 | -0.58 | -0.21 | 0.013 |
|  | Ischemic Stroke | -0.38 | -0.52 | -0.24 | 0.001 |
|  | Intracerebral | -0.30 | -0.50 | -0.13 | 0.016 |
|  | Hemorrhage |  |  |  |  |
|  | Subarachnoid | 0.85 | 0.74 | 0.95 | <0.00 |
|  | Hemorrhage |  |  |  |  |

DALYs, disability-adjusted life years.

### Stroke trends from 1990 to 2023 by sex

**Figure 3** shows declining age-standardized rates for both sexes, with a greater decline among women than among men for stroke incidence (−15.2% [95% UI: −18.5% to −11.6%]), deaths (−31.6% [95% UI: −49.0% to −5.3%]), and DALYs (−23.6% [95% UI: −41.0% to 2.7%]). Similar trends were observed for ischemic stroke and intracerebral hemorrhage. However, an increasing trend was observed for subarachnoid hemorrhage, with a more pronounced increase among men. Crude rates across all measures for both sexes increased since 1990, with men showing greater increase in stroke mortality (44.1% [95% UI: 5.0%–103.6%]) and DALY rates (46.8% [95% UI: 8.4%–104.4%]), whereas women showed greater increase in prevalence rates (40.2% [95% UI: 34.9%–45.9%]). (Supplementary Tables S1–4).

**Figure 3.**
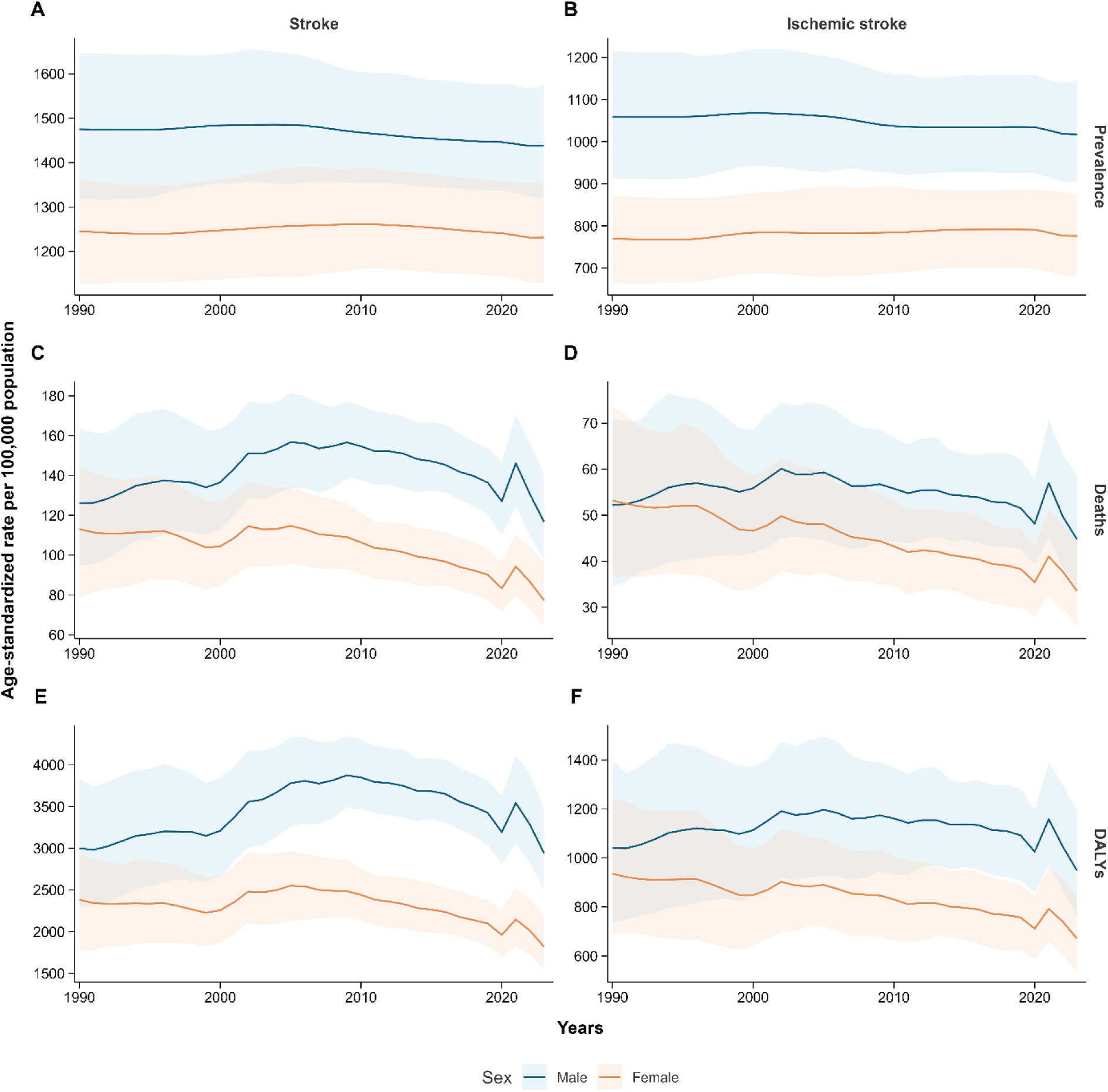
Temporal patterns of stroke in the Philippines by sex from 1990 to 2023

### Stroke trends from 1990 to 2023 by age

A declining trend among individuals aged 55 years was observed, as shown in Figure 3, and was most pronounced among individuals aged 80–84 years (incidence: −23.83% [95% UI: −17.27% to −28.82%]; mortality: −38.05% [95% UI: −22.61% to −50.67%]; DALYs: −36.10% [95% UI: −21.58% to −48.45%]). However, prevalence rates showed a notable increase among those aged 95 years and above (19.70% [95% UI: 35.13%–8.59%]). Younger adults (20–54 years), most notably in the 35–39-year age group, showed an increasing trend in stroke incidence, mortality, and DALYs rates, whereas prevalence rates increased in a wider age group (35–54 years). Similar patterns were also observed for ischemic stroke. Notably, subarachnoid hemorrhagic stroke rates exhibited a greater increase relative to other stroke subtypes, across a broader age group, and among older individuals. Increases in subarachnoid hemorrhagic stroke mortality and DALYs were most pronounced in the 15–19-year age group (mortality: 56.82% [95% UI: 184.84% to −21.21%]; DALY: 42.72% [133.89% to −19.42%]) (Supplemental Table S5–8).

**Figure 4.**
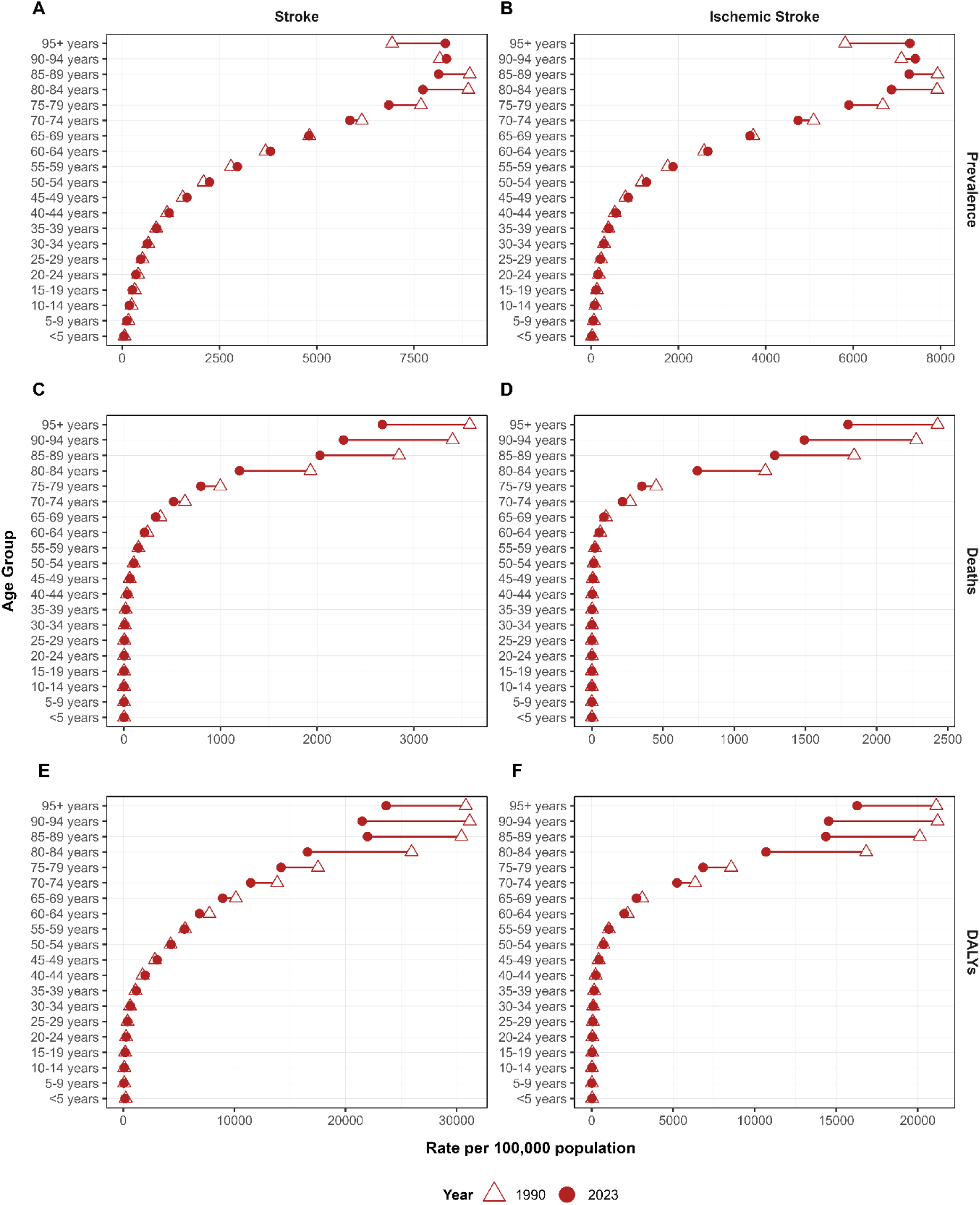
Temporal trends of stroke in the Philippines by age from 1990 to 2023

## Discussion

In this study, we report a 33-year comprehensive analysis of stroke burden in the Philippines (1990–2023) using the GBD 2023 dataset. In 2023, new and prevalent stroke cases accounted for 0.14% and 1.09%, respectively, of the estimated national population while stroke-related deaths and DALYs accounted for 10.90% and 6.26% of the total deaths and DALYs from all causes.^1^ Incident and prevalent cases were predominantly ischemic, consistent with previous findings.^5^ Although absolute numbers and crude rates have significantly increased since 1990, age-standardized rates declined—a trend projected to persist into the 2050s.^11^ This divergence suggests that, although the national stroke healthcare system has made progress in stroke control and management and individual-level risk has improved, the absolute burden continues to grow due to population growth and demographic aging—a pattern observed in both developing and developed countries.^12,13^ The Philippines is projected to have a significant older population^14^ which will amplify the stroke burden. The healthcare system must prepare for the structural demands of an aging society, including expanded neurorehabilitation services, long-term care, and integrated chronic disease management.^12^ Local government units (LGUs) should adopt and support the health component of the community-based inclusive development (CBID) programs initiated by the National Council on Disability Affairs to improve rehabilitation service delivery.^15^ Additionally, a strategic shift towards prospective bundled payments to cover the full continuum of stroke care from acute care to community-based rehabilitation can reduce catastrophic health expenditure and improve functional outcomes by incentivizing long-term follow-up.^16^

The provinces identified with the highest estimated age-standardized DALY and mortality rates are among the most underserved in terms of the availability of acute stroke-ready hospitals (ASRHs). These provinces belong to the Mindanao island group, where only 15 of the 91 certified ASRHs are located while the Visayas and Luzon island group have 13 and 63 hospitals, respectively. ^3^ This profound disproportionate distribution of facilities contributes to the persistent stroke burden. To improve stroke care in these high-burden provinces, telestroke hub spoke networks can be established to formally link non-certified institutions (spokes) to certified hubs in the region via 24/7 telestroke consultation and receive real-time guidance for acute stroke care.^15^

Metabolic risks, particularly high systolic blood pressure, were identified as the predominant risk factors contributing to stroke mortality and DALYs in the Philippines, whereas secondary risk factors included behavioral (tobacco smoking and alcohol use) and environmental (air pollution) risks, consistent with previous findings.^4,15^ This risk factor profile is highly consistent with the major drivers of stroke burden observed across lower-middle income countries (LMICs), where the burden is disproportionately high.^2,16^ Given the overwhelmingly attributable burden to potentially modifiable risk factors, a strategic shift toward primary prevention, supported by strong policy and system-level interventions, is imperative^15,17^ The fragmented implementation by LGUs of the national public health framework calls for the standardization and prioritization of hypertension management at the primary care level through the WHO HEARTS technical package. Additionally, consistent implementation of targeted population-wide interventions should be ensured to manage these risk factors and implement progressive community stroke screening and health education.

While fatal stroke burden (mortality and DALY) was higher among men in 2023, non-fatal burden (incidence and prevalence) was higher among women due to longer life expectancy and stroke onset at a later age.^2,18^ Sex-specific analysis revealed a pronounced epidemiologic divergence: men consistently exhibited higher age-standardized burden than that exhibited by women, however, women achieved substantially greater stroke burden reduction over the same period. The progress differences between sexes were driven by specific trends in ischemic and hemorrhagic stroke subtypes. Women generally achieved greater reductions in the burden of ischemic stroke and intracerebral hemorrhage, whereas subarachnoid hemorrhage exhibited an increasing age-standardized burden in both sexes. These differences are attributable to biological differences and varying exposures to established risk factors. In premenopausal women, estrogen offers protective effects against stroke, and the loss of this effect after menopause contributes to the increase in stroke incidence among older women^18,19^ Hormonal influences, together with variations in coagulation profiles, may contribute to an elevated risk of hemorrhagic stroke in women.^20^ Smoking and alcohol consumption are consistently identified as important risk factors for men.^12,21^

Age-specific analysis indicated that stroke- and ischemic stroke-related DALYs were predominately concentrated among older adults, whereas hemorrhagic stroke-related DALYs were disproportionately higher among younger adults than older adults. Furthermore, although individuals aged 65 years and above bore the greatest stroke burden, they achieved substantial reductions in stroke incidence and stroke-related deaths. In contrast, both fatal and non-fatal burden increased markedly among younger adults (15–54 years). This trend among individuals in their prime working years has significant implications for disability, health care costs, and potential economic productivity^15,22^ Future stroke prevention strategies must specifically target strokes in young and middle-aged groups. Integration of cardiovascular assessment into routine prenatal and postnatal care for women of reproductive age has previously been recommended^23^ Additionally, policies that mandate annual physical examinations among all Philippine workers should be considered, and the examination should include a cardiovascular risk score.^3^

These divergences reflect a broader pattern observed across the Southeast Asia and Western Pacific (WP) regions. The decline in age-standardized incidence rates remains modest in the Philippines compared with high-income regional counterparts such as South Korea.^1,25^ Patterns in stroke subtypes were also consistent, where these two regions accounted for majority of the global DALYs lost to intracerebral hemorrhage.^25^ The disproportionate increasing risk among younger individuals was also observed in China and other LMICs in the WP region that similarly struggle with controlling modifiable risk factors.^2,24,26^ The persistent high stroke burden in the Philippines, relative to its regional counterparts, is exacerbated by the prevailing gaps in the stroke care system. The density of critical diagnostic equipment, such as computed tomography scanners, in the Philippines is significantly lower than that in Brunei, Thailand, and Singapore. The Philippines’ devolved healthcare system, which has led to substantial regional disparities, differs from the centralized organized system in South Korea. Philippines health financing mainly relies on out-of-pocket spending, accounting for a substantial amount of stroke cost, a barrier that is less pronounced in high-income neighboring countries with more comprehensive universal coverage.^3^ Drawing from these comparisons with neighboring countries, the Philippines is uniquely challenged by earlier onset of stroke and disproportionate distribution of stroke care.

To the best of our knowledge, this is the first study to comprehensively analyze the temporal trends in the burden of stroke and its pathological subtypes in the Philippines using the GBD database. However, several limitations inherent to the GBD methodology and the local health landscape should be acknowledged and considered when interpreting these results. As a secondary analysis, the validity of our findings is linked to the quality and completeness of primary Philippine data sources, such as vital registrations and hospital records. Using such data may not capture the full burden of stroke since approximately one-third of the population dies without medical assistance.^3^ This is further compounded by the disproportionate geographical distribution of stroke-ready hospitals, likely contributing to under-reporting and misclassification of stroke subtypes in certain provinces and to the exclusion of hospital records with extremely low number of cases from the analysis.^1^ These gaps necessitated the interpolation of missing provincial-level estimates. While such estimations ensure a complete time series, they result in wider UIs among data-poor regions, indicating that the actual burden in these provinces may be subject to greater measurement error. Consequently, while our results provide a robust perspective on national trends, subnational disparities should be interpreted as conservative estimates. With the urgent need for continued, systematic surveillance of trends, this study highlights the need to enforce comprehensive reporting to the national stroke registry by all health facilities to capture the full burden of stroke, enable systematic surveillance, and ensure evidence-based policy and clinical decisions.

## Conclusion

Although age-standardized stroke burden decreased, absolute number and crude rates in the Philippines showed increasing trends from 1990 to 2023. Ischemic stroke remains the most prevalent stroke subtype; however, hemorrhagic strokes account for a larger proportion of fatal burdens. The study findings highlight the need for strengthened and widespread acute and post-stroke interventions, especially among male and younger adults. Furthermore, a paradigm shift toward aggressive prevention of modifiable risk factors for stroke is needed.

## Data Availability

This study utilized publicly available data from the Global Burden of Disease (GBD) 2023 study, which provides estimates of stroke burden and risk factors across countries and subnational locations. All the data used in this analysis can be accessed through the Institute for Health Metrics and Evaluation GBD Results Tool and GBD Compare visualization platform at http://ghdx.healthdata.org/gbd-results-tool and http://vizhub.healthdata.org/gbd-compare, respectively

http://ghdx.healthdata.org/gbd-results-tool

## Acknowledgment

The authors would like to recognize the works by the Global Disease and Injury and Risk Factor Study 2023 collaborators. We also would like to thank Editage (www.editage.jp) for English language editing.

